# Association between comorbidities and death from COVID-19 in different age groups

**DOI:** 10.1101/2021.04.12.21255365

**Authors:** Pedro Emanuel Fleitas, María Cristina Almazán, Sabrina Daniela Cortez, Jorge Augusto Paz, Rubén Oscar Cimino, Alejandro Javier Krolewiecki

**Affiliations:** Instituto de Investigaciones de Enfermedades Tropicales (IIET-CONICET), Universidad Nacional de Salta. Alvarado 751. San Ramón de la Nueva Orán, Salta, Argentina. ZC: 4530; bCátedra de Química Biológica, Facultad de Ciencias Naturales, Universidad Nacional de Salta. Avenida Bolivia 5150. Salta, Argentina. ZC: 4400; Instituto de Estudios Laborales y del Desarrollo Económico (IELDE), Consejo Nacional de Investigaciones Científicas y Técnicas (CONICET), Universidad Nacional de Salta. Avenida Bolivia 5150. Salta, Argentina. ZC: 4400

**Author notes:** **Corresponding autor**: PhD. Pedro E. Fleitas, Mail, Work address: Instituto de Investigaciones de Enfermedades Tropicales (IIET). Alvarado 751, San Ramón de la Nueva Orán, Salta, Argentina. Zip Code: 4530.

**Keywords:** COVID-19, Comorbidities, Fatality rate, Score

## Abstract

**Background:** This new COVID-19 pandemic challenges health systems around the world; therefore, it is extremely important to determine which patients with COVID-19 can evolve to more severe outcomes. Accordingly, we decided to assess the role that comorbidities play in death from COVID-19.

**Methods:** Two age groups (<60 and ≥ 60 years) were defined for analysis. Decision trees were made to identify which comorbidities had the highest fatality rate (FR). Multiple logistic regressions were performed to measure the association between comorbidities and death.

**Results:** A significant difference was found between the FR of <60 group and ≥ 60 group. The most frequent comorbidity were cardiac diseases and diabetes. The combination of comorbidities with the highest FR was diabetes with kidney disease. Combinations of more than two comorbidities presented higher FR. The comorbidities had higher Odd ratios in the younger group than in the older group.

**Conclusions:** Comorbidities seem to play a greater role in death from COVID-19 in the younger group, while in the> 60 group; age seems to be the most important factor. We assigned a score to the comorbidities and their combinations for both age groups to help the health personnel make decisions.

## Introduction

From the first case of COVID-19 that was reported in the city of Wuhan, China in December 2019 until the beginning of November 2020, 46 million infected people have been reported worldwide.^1^ Of these 46 million, 2.6% have died. ^1^ This new pandemic challenges health systems around the world and carries important ethical questions, especially under the need of rationing health care in the context of scarce resources and crisis capacity. ^2^ For this reason, it is extremely important to determine which patients with COVID-19 can evolve to more severe outcomes, including death. Age, male gender, and ethnicity have all been reported to be associated with severe COVID-19 outcomes.^3,4^ Also, comorbidities such as diabetes, kidney, respiratory, and cardiovascular diseases have been reported as risk factors for severe outcomes by COVID-19. ^3,5^ Based on this, we decided to assess the role that comorbidities play in death from COVID-19 in a large database from Brazil that includes recovered and deceased patients, to provide valuable information for designing strategies for case management.

## Material and methods

### Data source

The data file was downloaded from the website of the Brazilian Ministry of Health on August 1, 2020. ^6^ The data comes from the e-SUS NOTIFICA system, which was developed to register suspected cases of COVID-19 in Brazil, and contains patient information such as place of residence, clinical, demographic, and epidemiological characteristics. An *ad-hoc* database was built for this project with all laboratory confirmed positive cases (RT-PCR or serology) or clinical-epidemiological survey containing information on date of onset of symptoms, date of the diagnostic test, age, sex, and disease evolution. Patients with missing data were excluded from the database. We did not impose any further exclusion criteria to limit selection bias.

### Study design

Two age-group were defined for the analysis with those <60 or ≥ 60 years-old. Decision trees were built to order the comorbidities according to their frequency and the proportion of deaths from COVID-19 reported. Then, the correlation of the absence/presence of the following comorbidities: diabetes, immunosuppression, kidney, respiratory, and cardiovascular disease with death from COVID-19 was analyzed. For this, a multiple logistic regression was performed to identify which variables were associated with death from COVID-19. Also, the probability obtained from logistic regressions (probability of dying when infected by COVID-19 if the comorbidities were present) was used to calculate the area under the receiver-operator characteristic curve (AUC). The AUC was used as a measure of the ability of the comorbidities model to predict death from COVID-19. Finally, the values of probability obtained from logistic regressions were used to assign a score to the comorbidities and the combinations of them. All statistical analyses were carried with RStudio version 1.3.1093 (RStudio: Integrated Development for R. RStudio, PBC, Boston, MA). Analyses with p-values <0.05 for a level of significance of 95% were considered statistically significant. Figures were done with GraphPad Prime 9 Trial version (GraphPad Software Inc., San Diego, CA).

## Results

### Data description

The whole database includes over 8 million cases recorded in the 27 states of Brazil from January 1 to August 1 2020, 549733 were included in the analysis (Fig. 1). A summary of the comorbidities registered in all the patients can be observed in Table S1 of supplementary material. In our database, median age was 38 years old (IQR: 30-51); 46% were males and 54% females. It was found that 2.1% of patients died while 97.9% were recovered. A statistically significant difference was found between the fatality rate of <60 group (0.7%, n= 457557) and ≥ 60 group (11.3% n= 74176) (p<0.01) (Fig. 2). Also, the median time between the onset of symptoms and death was 20 days (IQR: 12-33), and no differences were found between age groups.

**Figure 1:**
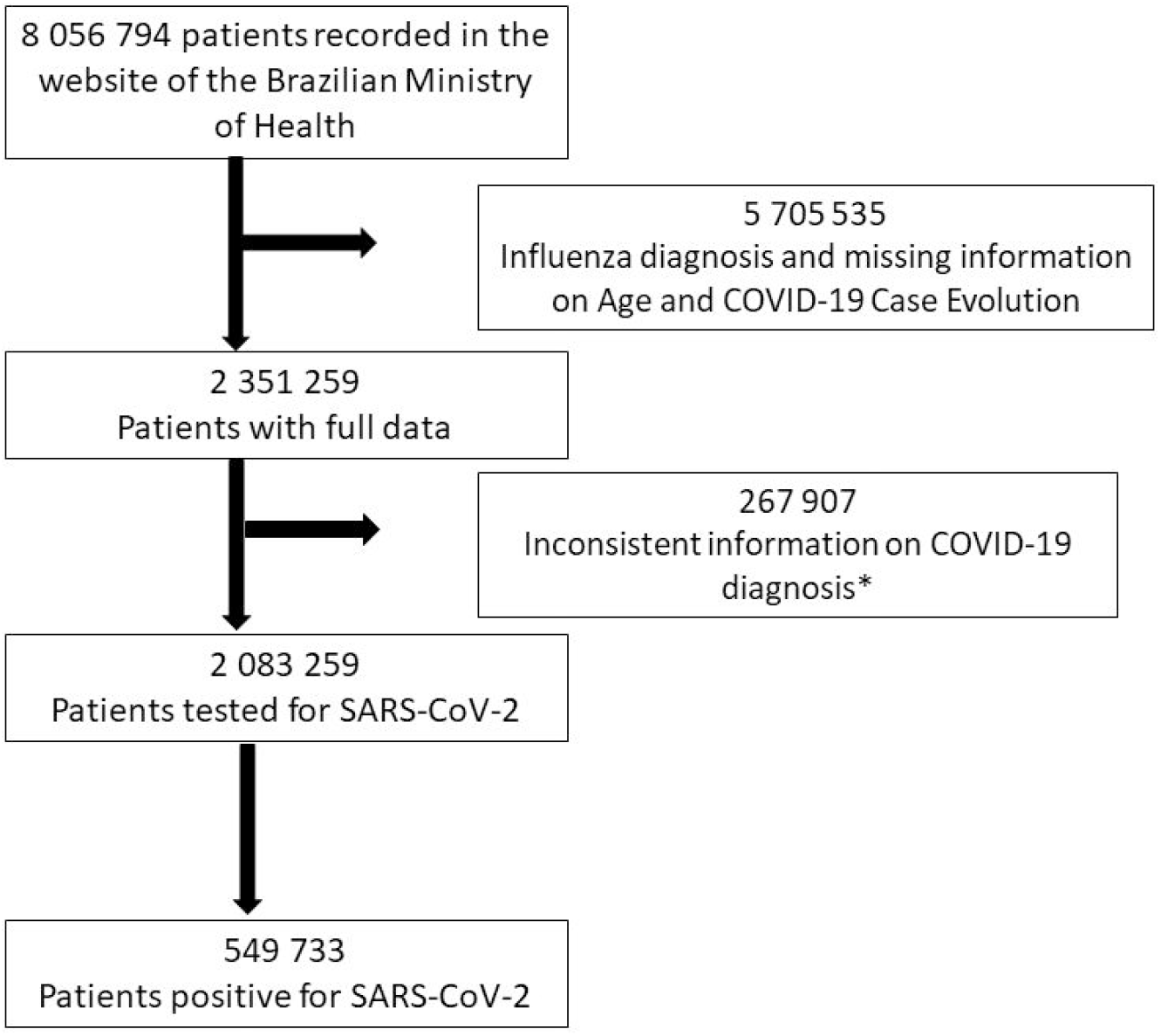
Flowchart of data used in this study. *According to Guidelines for Case Notification of Influenza Syndrome suspected of COVID-19.^7^

**Figure 2:**
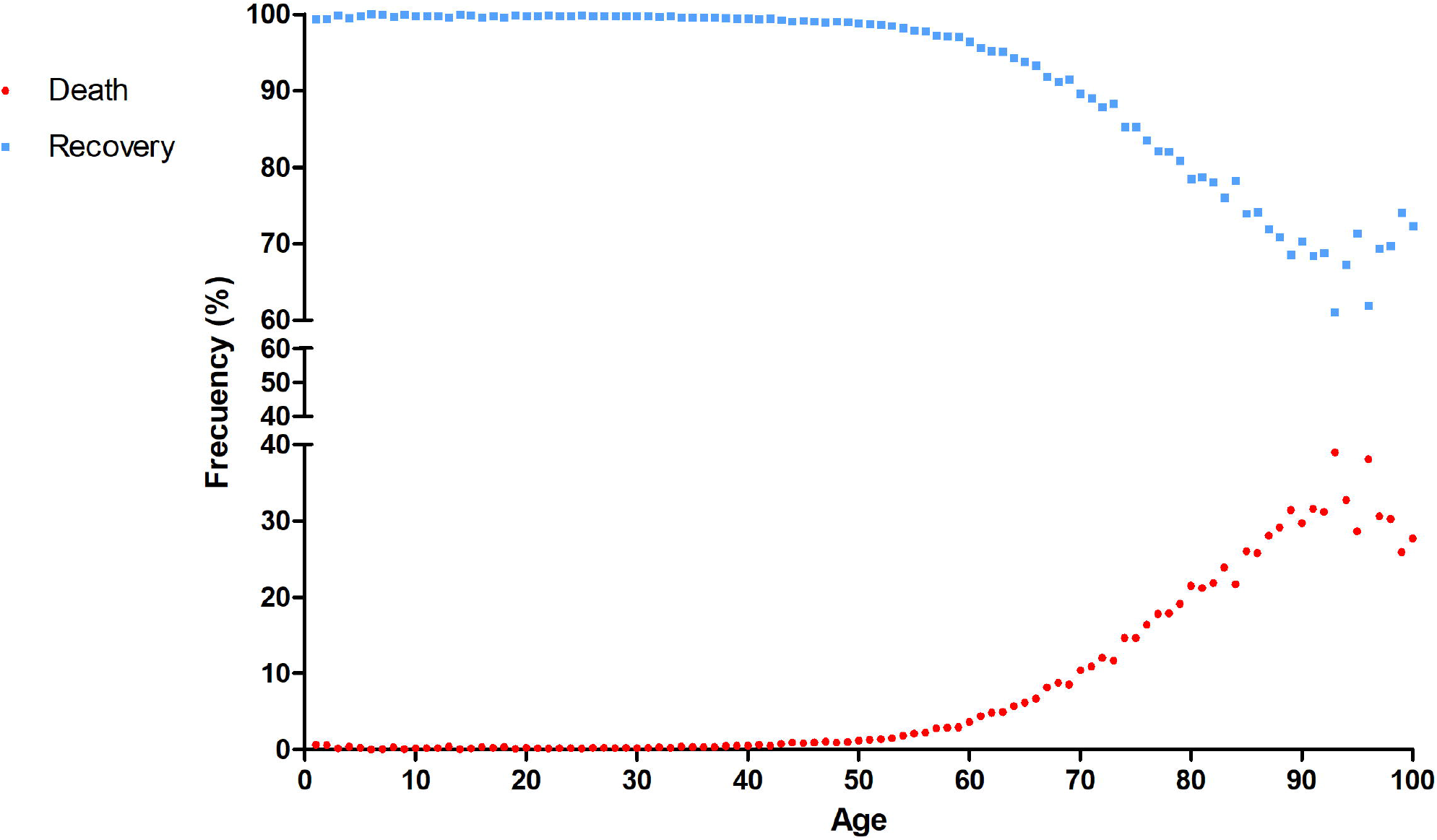
Frequency of recovery and death from COVID-19, according to the patient’s age.

Five comorbidities were recorded in the data file: cardiovascular disease, immunosuppression, kidney disease, respiratory disease, and diabetes. Among 87.6% of the cases, no comorbidities were registered; meanwhile, the most frequent comorbidity registered in the database was cardiovascular disease (7.2% (39542/549733)), followed by diabetes (4.3% (23966/549733)), respiratory disease (2.0% (11035/549733)), immunosuppression (0.9% (4934/549733)) and kidney disease (0.5% (2684/549733)). In addition, It was observed that in the <60 group, cardiac disease (Frequency= 4.3%) and diabetes (Frequency= 2.6%) presented a fatality rate of 1.6% and 0.5% (Fig. 3a). In addition, the highest fatality rate (19%) was found in individuals with diabetes and kidney disease (Fig. 3a). In the ≥60 group the most frequent comorbidities were also diabetes (Frequency =15%, fatality rate =19%) and cardiovascular disease, without diabetes, (Frequency=12.4%, fatality rate =13%). At the same time, the combination of comorbidities with the highest fatality rate was diabetes with kidney disease (fatality rate= 35%) and diabetes with kidney and respiratory diseases (fatality rate= 47%) (Fig. 3b).

**Figure 3:**
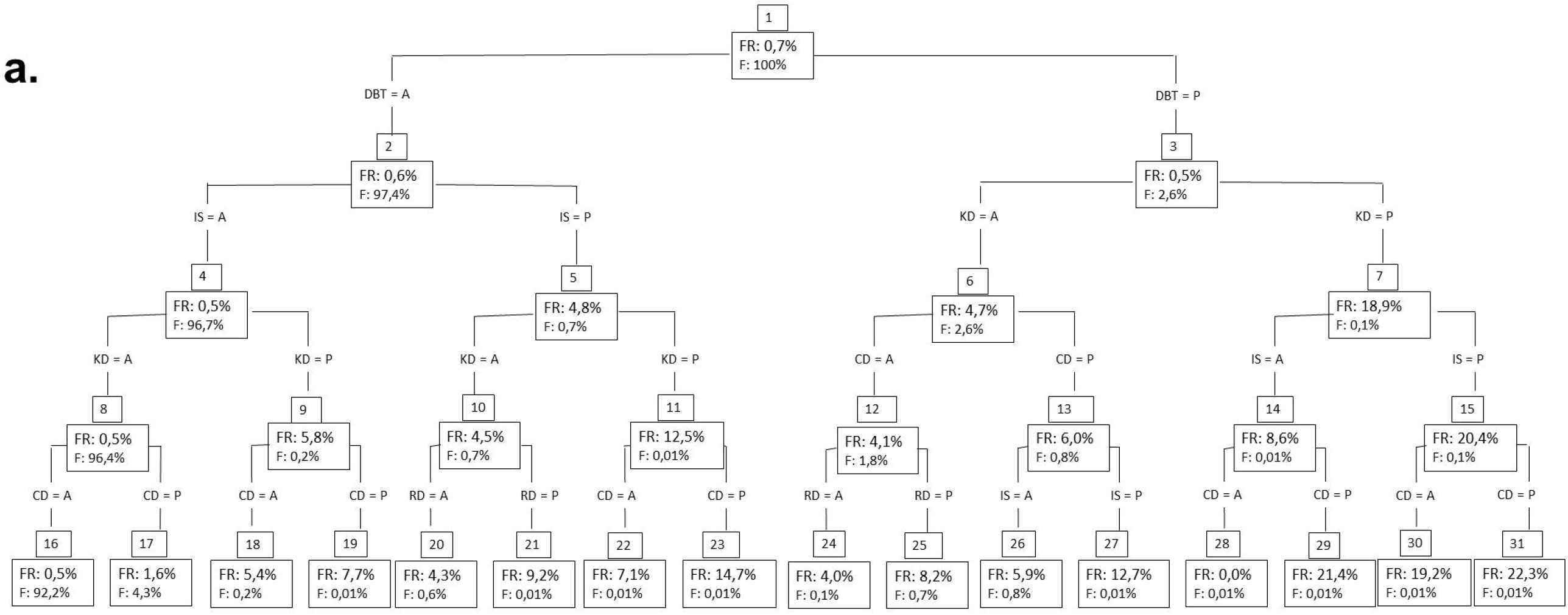
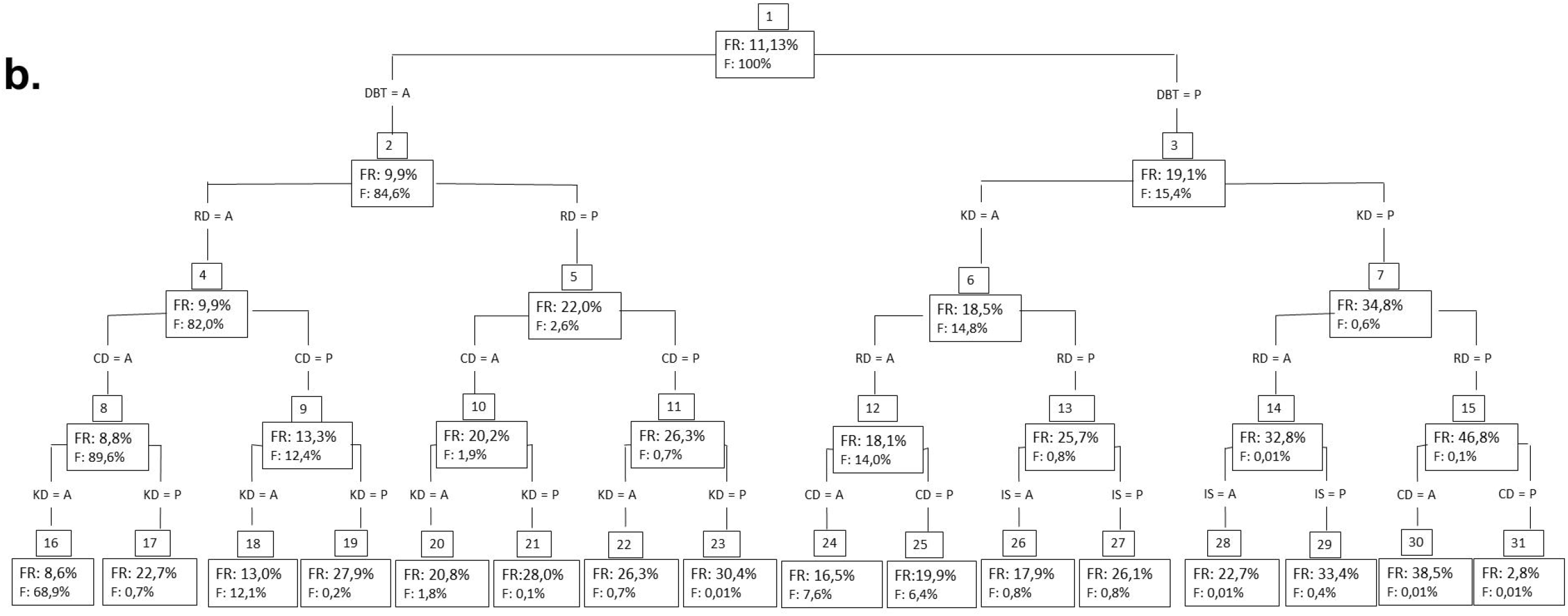
Tree diagrams representing the frequency of the comorbidities and the fatality rate. **a**. <60 group (n=457557). **b**. ≥ 60 group (n=74176). In each node, its number is represented in the top, and inside the fatality rate (MR) and the percentage frequency (F). DBT= Diabetes, IS = immunosuppression, KD = kidney disease, RD = respiratory disease, CD= Cardiovascular disease, P= comorbidity present and A= comorbidity absent.

### Correlation among comorbidities and death from COVID-19

For the <60 group and ≥ 60 groups, the values of AUC=0.66 and AUC=0.61 were obtained, respectively. As Fig. 4 shows, the comorbidities had higher ORs in the <60 group than in the ≥ 60 group. In this first group, diabetes, kidney disease, and immunosuppression had the highest ORs; whereas in the ≥60 group, the comorbidities with the highest ORs were kidney and respiratory diseases (Fig. 4).

**Figure 4:**
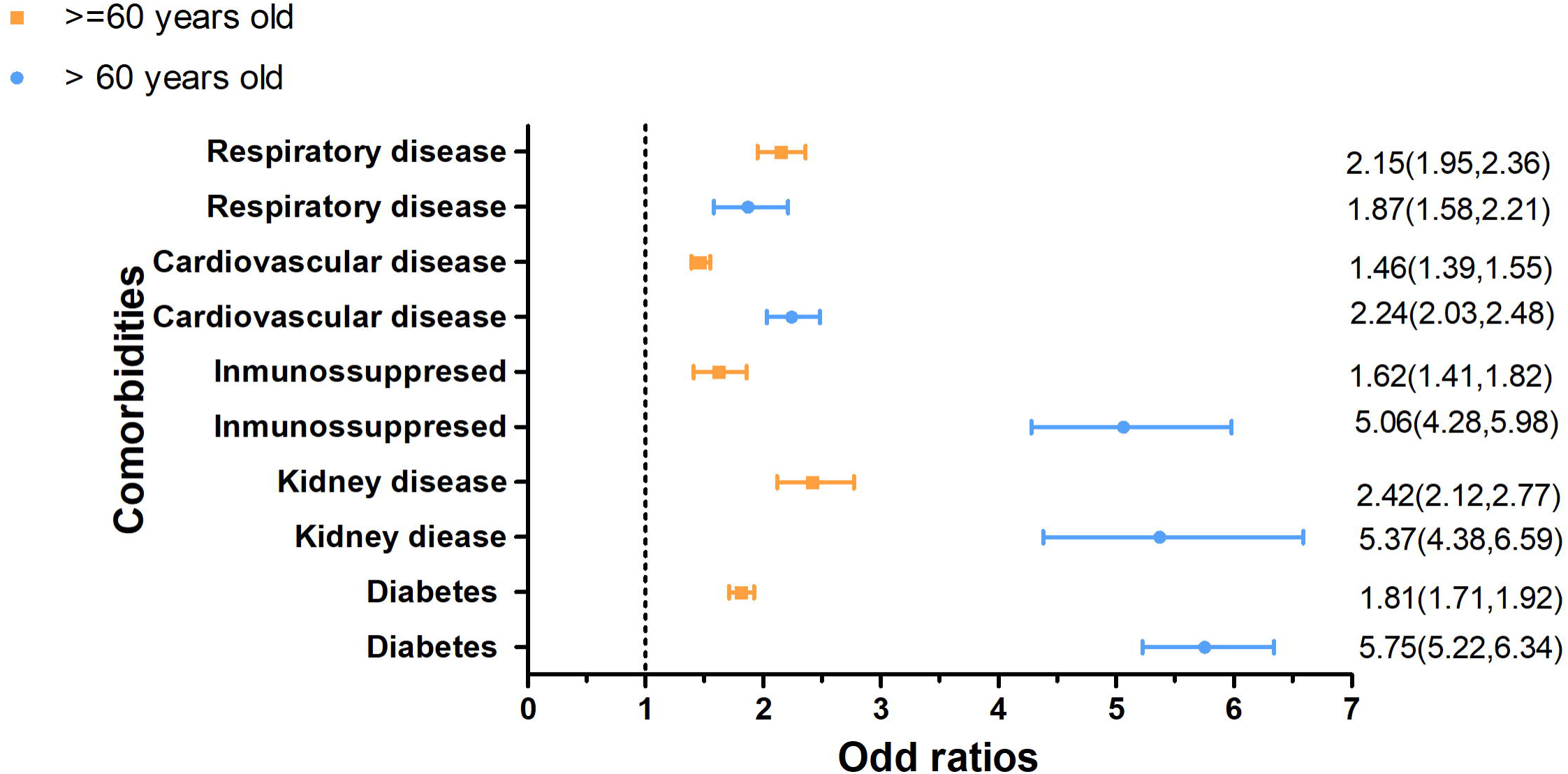
Odds ratios for comorbidities in the <60 group and ≥60 age group.

The logistic regression allowed us to assign a score to the comorbidities and their combinations, from one (lower probability of a severe outcome) to eight (high probability of a severe outcome) for both age groups. These scores were ordered into three categories of low, medium, and high risk, based on the incidence of death from COVID-19. It can be observed that the highest score is found with the combination of diabetes and kidney disease or the combination of 3 or more comorbidities.

**Table 1:** Score assigned to the different comorbidities based on the probability of death when infected by COVID-19 (multivariate logistic regression) and the fatality rate.

| <60 group |  |  |  |
| --- | --- | --- | --- |
| Score range |  | Comorbidities | Assigned score |
| **Low Score** | Probability<br><0.02<br>Fatality rate<br>=0.5<br>n=458076 | Cardiovascular disease | 1 |
|  |  | Respiratory disease | 1 |
| **Medium Score** | Probability<br>=0.02-0.04<br>Fatality rate<br>=4.0<br>n=12352 | Cardiovascular disease +<br>Respiratory disease | 2 |
|  |  | Immunosuppression | 2.5 |
|  |  | Kidney disease | 3 |
|  |  | Diabetes | 3.5 |
| **High Score** | Probability<br>≥0.05<br>Fatality rate<br>=7.0<br>n=5129 | Immunosuppression +<br>Respiratory disease | 4 |
|  |  | Kidney disease + Respiratory<br>disease | 5 |
|  |  | Diabetes + Respiratory<br>disease | 5.5 |
|  |  | Immunosuppression +<br>Cardiovascular disease | 6 |
|  |  | Cardiovascular disease + Renal disease | 6.5 |
|  |  | Diabetes + cardiovascular disease | 7 |
|  |  | Immunosuppression + Kidney disease | 7.5 |
|  |  | Diabetes + Kidney disease or ≥ 3 comorbidities | 8 |

| ≥ 60 group |  |  |  |
| --- | --- | --- | --- |
| Score range |  | Comorbidities | Assigned score |
| **Low Score** | Probability <0.17<br>Fatality rate =9.9<br>n=65466 | Cardiovascular disease | 1 |
|  |  | Immunosuppression | 1 |
|  |  | Diabetes | 1 |
| **Medium Score** | Probability 0.17-0.29<br>Fatality rate= 20.7<br>n=7801 | Respiratory disease | 2 |
|  |  | Immunosuppression + Cardiovascular disease | 2.5 |
|  |  | Kidney disease | 3 |
|  |  | Cardiovascular disease + diabetes | 3.5 |
|  |  | Diabetes + Immunosuppression | 4 |
|  |  | Cardiovascular disease + Respiratory disease | 4.5 |
|  |  | Immunosuppression + Respiratory disease | 5 |
|  |  | Cardiovascular disease + Kidney disease | 5.5 |
|  |  | Diabetes + Respiratory disease | 6 |
|  |  | Immunosuppression + Kidney disease | 6.5 |
| **High Score** | Probability $\geq 0.29$<br>Fatality rate = 30.1<br>n=909 | Renal disease + Respiratory disease | 7 |
| | | Diabetes + Kidney disease or $\geq 3$ comorbidities | 8 |

## Discussion

Since the beginning of the COVID-19 pandemic in December 2019, the reported case fatality rate has varied. Early estimates from small cohorts of hospitalized patients indicated a fatality rate of 15%; later and with more data, the percentage of deaths fell from 11% to 4.3% and finally to 3.4%.^8^ In addition, 81% of patients with COVID-19 have a mild illness and never require hospitalization.^9^ Therefore, for a better understanding of the causes that lead to severe results of COVID-19, in this retrospective cohort study, we analyzed risk factors for death in individuals with COVID-19 in a large database from Brazil with a fatality rate of 2.1%.

Older age has been reported by several authors as an important factor associated with the severity of the disease or fatality in patients with COVID-19.^4,5,10^ In accordance with this, we found a higher fatality rate in patients older than 60 years of age.

It is important to highlight the relevance of age-stratified analyses when examining the association between comorbidities and fatality with COVID-19.^5^ In our study, the odd ratio (OR) values for comorbidities varied depending on age group. In the <60 group, both higher ORs and a higher AUC value were observed compared to those obtained for the ≥ 60 group. This suggests that the comorbidities considered are able to better explain the fatality from COVID-19 in the <60 group than in the ≥ 60 group. Therefore, the comorbidities seem to have an important impact on death in the <60 group, while in the ≥ 60 group the comorbidities might play a minor role in death from COVID-19. This suggests that the main risk factor in the older group is age itself, which is consistent with previous findings for Brazilians patients.^3^

Diabetes was the most frequent comorbidity and had the highest OR value in the <60. Patients who presented diabetes as comorbidity, reached a fatality rate of 0.5% and 19% in the <60 group and ≥ 60 group, respectively (Fig. 2). Worth mentioning, Brazil is one of the countries with more diabetes cases in the world, being the estimated prevalence of 11.9 % (95 % CI 7.7–17.8).^11^ The evolution of COVID-19 has shown to be more severe in patients with diabetes and metabolic dysfunction.^12^ Recent data suggests that COVID-19 could precipitate acute metabolic complications of diabetes, such as diabetic ketoacidosis and hyperglycemia. The potential mechanisms that may increase susceptibility to severe COVID-19 in patients with diabetes include: 1) higher affinity cell binding and effective entry of the virus, 2) decreased viral clearance, 3) decreased T cell function, 4) increased susceptibility to hyperinflammation and cytokine storm syndrome, and 5) presence of cardiovascular diseases.^13^ This becomes more relevant when considering that the most frequent combination of comorbidities included diabetes and cardiovascular disease, with a fatality rate of 6% and 20% in the <60 group and≥ 60 group, respectively.

Another important risk factor is kidney disease. In the ≥ 60 group, this comorbidity together with respiratory disease presented the highest ORs. Moreover, the combination of diabetes with kidney disease caused the highest fatality rates. Since diabetes is the main cause of chronic kidney disease,^14^ patients with these two comorbidities can represent diabetic patients in a more severe stage of the disease or without adequate medical care. Interestingly, the human kidney has been reported to be a target for SARS-CoV-2^15^; however, it remains controversial whether SARS-CoV-2 causes acute kidney injury.^16^

There is a wide spectrum of immunosuppressed patients, including patients with cancer, solid organ transplant recipients, those taking antirheumatic drugs, primary immunodeficiency, and HIV infection. We found that in both age groups, immunosuppression was a risk factor associated with death from COVID-19. However, this should be interpreted with caution since the database did not allow to determine precisely the underlying cause of the immunosuppression and because there is controversy about the role immunosuppression in COVID-19. Previous research indicates that patients with cancer and solid organ transplant recipients may have a higher risk of developing more severe COVID-19.^17,18^ Nonetheless, immunosuppressed adults with no further comorbidities might not be likely to have a severe clinical outcome.^18^ This contradiction may be because the host’s immune response is an important factor contributing to the severity of COVID-19, through a dysregulation of innate immunity or an excessive inflammatory response.^19^ Thus, immunosuppression, as a sole comorbidity, probably does not determine a severe outcome.

As has been shown in this study, there are comorbidities that increase the probability of severe COVID-19. Therefore, they should be considered in case management. Concerning this, we assigned a score to each comorbidity to provide a possible order of priority. Based on the analysis of this large database, there are small subgroups of cases that carry an exceedingly high risk of death due to COVID-19.

It is important to understand the limitations of this study when interpreting these results. First, possible selection biases cannot be ruled out due to the reduction of the data file. Second, the data come from all the states of Brazil, but might not be representative of the entire population, and the generalizability of the results beyond this cohort is unclear. However, it is worth noting the high number of cases analyzed and that the proportion of the comorbidities in the database (n=549733) is similar to in the original data file (n=8056794).

In conclusion, we demonstrate that diabetes, kidney disease, respiratory disease, cardiovascular disease, and immunosuppression are risk factors related to death from COVID-19, where age seems to be a determining factor. The combination of comorbidities increases the risk, which must be considered and analyzed for clinical decisions.

## Data Availability

All data used in this work are freely available and provided by the Brazilian Ministry of Health.

https://opendatasus.saude.gov.br/dataset/casos-nacionais

## Ethics approval

The material used was freely available on the website of the Brazilian Ministry of Public Health, so it was not necessary to have ethical approval.

## Supplementary materials

Supplementary material associated with this article can be found in the online version.

