## Supplemental Table 1 for "Association between comorbidities and death from COVID-19 in different age groups"

**Supplementary Material.**

Table S1: Summary of the data file.

| **State** | **n** | ***Positive for SARS-CoV-2** | ****Deaths** | **Cardiovascular disease** | **Diabetes** | **Respiratory disease** | **Immunosuppression** | **Kidney disease** | **Median age** |
| --- | --- | --- | --- | --- | --- | --- | --- | --- | --- |
| Acre | 55274 | 12147 | 420 | 3116 | 1683 | 1301 | 846 | 417 | 38 |
| Alagoas | 215420 | 37679 | 1341 | 12146 | 9625 | 3244 | 2133 | 838 | 38 |
| Amazonas | 236334 | 38403 | 237 | 9177 | 6254 | 3815 | 1382 | 532 | 39 |
| Amapá | 67405 | 22891 | 523 | 2843 | 1723 | 1246 | 438 | 233 | 38 |
| Bahia | 681031 | 138711 | 3612 | 29927 | 21024 | 11781 | 5301 | 3685 | 38 |
| Ceara | 521124 | 111413 | 2740 | 13507 | 9549 | 4661 | 1409 | 869 | 39 |
| Distrito Federal | 190132 | 24057 | 204 | 11656 | 7792 | 5772 | 2753 | 1335 | 37 |
| Espírito Santo | 15014 | 879 | 8 | 634 | 488 | 269 | 151 | 195 | 39 |
| Goiás | 315814 | 71673 | 433 | 11559 | 7744 | 6899 | 2688 | 1502 | 37 |
| Maranhão | 40161 | 18314 | 559 | 2006 | 1379 | 622 | 199 | 170 | 38 |
| Minas Gerais | 922422 | 52532 | 1071 | 42449 | 26579 | 29646 | 7955 | 3576 | 36 |
| Mato Grosso Sul | 137787 | 26758 | 30 | 5901 | 3841 | 3100 | 1204 | 518 | 36 |
| Mato Grosso | 131942 | 7405 | 214 | 5795 | 4499 | 2038 | 847 | 446 | 35 |
| Pará | 311277 | 53106 | 1205 | 14687 | 10014 | 6161 | 2243 | 1454 | 38 |
| Paraíba | 333512 | 53542 | 531 | 17986 | 10298 | 5853 | 2361 | 987 | 38 |
| Pernambuco | 223303 | 47905 | 300 | 8842 | 6138 | 4018 | 1423 | 947 | 38 |
| Piauí | 176311 | 21463 | 300 | 8038 | 4782 | 2831 | 1195 | 511 | 37 |
| Paraná | 287334 | 3726 | 129 | 8208 | 14912 | 7889 | 823 | 1348 | 38 |
| Rio de Janeiro | 779714 | 55642 | 1745 | 64814 | 33806 | 23898 | 7031 | 3903 | 39 |
| Rio Grande do Norte | 242013 | 16443 | 655 | 25101 | 14565 | 7835 | 3055 | 1727 | 39 |
| Rondônia | 162576 | 40398 | 930 | 7206 | 4544 | 3474 | 1618 | 1015 | 38 |
| Roraima | 78333 | 6313 | 87 | 4983 | 2740 | 1673 | 585 | 532 | 37 |
| Rio Grande do Sul | 540341 | 70830 | 251 | 27625 | 14272 | 17549 | 5330 | 2156 | 37 |
| Santa Catarina | 316908 | 45413 | 395 | 10906 | 7205 | 6883 | 2626 | 847 | 37 |
| Sergipe | 121272 | 12198 | 8 | 4880 | 3345 | 1674 | 692 | 443 | 37 |
| São Paulo | 868859 | 129034 | 866 | 55202 | 36641 | 29162 | 8700 | 4371 | 39 |
| Tocantins | 85181 | 16344 | 268 | 3126 | 1995 | 1307 | 646 | 250 | 35 |
| Total | 8056794 | 1135219 | 19062 | 412320 | 267437 | 194601 | 65634 | 34807 | 38 |

* Positive through laboratory diagnostic and clinical epidemiological survey

** Total deaths, including negative cases for COVID-19
